# Biomarkers of pesticide exposure predict Depressive Symptoms in rural and urban workers of the Maule Region, Chile

**DOI:** 10.1101/2025.01.30.25321387

**Authors:** Boris Lucero, María Teresa Muñoz-Quezada, Andrés Canales-Johnson, Cynthia Carrasco, María Victoria Rodríguez, Tannelle Reid, Pablo Méndez, Jandy Adonis, Tristan A. Bekinschtein

## Abstract

**Background:** Agricultural workers are routinely exposed to pesticides, which are associated with a range of adverse health outcomes, including potential impacts on mental health such as depressive symptoms. In the Maule region of Chile, an area characterized by intensive pesticide use, this study investigates the association between exposure to organophosphate (OP) pesticides and the prevalence of depressive symptoms among agricultural workers.

**Methods:** We conducted a longitudinal study with 78 participants, comprising agricultural workers occupationally exposed to pesticides and nearby urban non-agricultural workers. Pesticide exposure was assessed through six dialkyl phosphate (DAP) metabolites in four repeated urine samples over two years. Depressive symptoms were evaluated using the CES-D scale and affect using the PANAS scale. Sociodemographic, health, and exposure-related variables were included as covariates. Generalized Estimating Equations (GEE) were employed to investigate dose-response relationships between pesticide exposure and depressive symptoms.

**Results:** No reliable differences in urinary DAP, DEAP, and DMP metabolite concentrations were observed directly when testing between rural and urban workers, however, dose-response analyses revealed that cumulative exposure to diethyl alkyl phosphates (DEAPs) was associated with depressive symptoms (CI=0.0003–0.0127, p = 0.040). Over time, positive affect increased among rural workers, whereas urban workers reported significantly lower negative affect in 2023. Further analyses showed problematic alcohol consumption (CI=0.018–3.707, p = 0.048) as a predictor of depressive symptoms.

**Conclusions:** Pesticide exposure extends beyond occupational settings to include environmental sources or food residues, as shown by comparable exposure levels in rural and urban participants and consistent measurements over two years. Chronic exposure to DEAPs was predictive of depressive symptoms, emphasizing the importance of biomarkers for assessing cumulative exposure. Public health interventions, stricter regulations, and awareness programs are needed to address the mental health risks of pesticide exposure. Further research should explore genetic and environmental factors influencing susceptibility to mental health.

## 1. Introduction

Pesticides are widely used in agriculture to control pests and protect crops.^1^ While these agrochemicals were designed to enhance agricultural productivity, growing evidence highlights their harmful effects on human health.^2,3^ Prolonged exposure to neurotoxic agricultural chemicals has been associated with mental health disorders, neurobehavioral changes, and other chronic conditions.^4^

Climate change has driven increased pesticide use to combat rising pest and disease pressures on crops.^5^ This has heightened exposure among agricultural workers, worsening associated health risks.^6^ Despite regulations to limit pesticide residues and reduce occupational and environmental exposure, significant health risks for agricultural workers remain.^7^

Research has shown a strong association between pesticide exposure and various health issues, including mental disorders like depression.^3,4,8,9^ Notably, pesticide exposure is recognized as a risk factor for suicide, particularly in rural communities and among agricultural workers.^10,11,12^ Recent studies indicate a strong link between pesticide exposure and increased depressive symptoms among agricultural workers.^13,14^ Research in South Korea^15^ reported a significant association between pesticide exposure and depression in farmers, with acute pesticide poisoning markedly increasing the risk (OR = 5.83; 95% CI, 1.80–18.86). Even chronic low-level exposure was associated with a higher likelihood of depression (OR = 1.67; 95% CI, 1.05–2.66). These findings highlight both acute and chronic pesticide exposures as significant risk factors for depression in agricultural populations.

Despite growing evidence, there is a lack of longitudinal studies using exposure biomarkers and validated assessments for depressive symptoms, particularly among agricultural workers in Latin America.^8^ A recent meta-analysis^3^ reported a significant association between pesticide poisoning and depression, with an OR of 2.94 (95% CI, 1.79–4.83, *p* < 0.001), indicating that individuals with acute pesticide poisoning are nearly three times more likely to develop depression. However, the link between chronic pesticide exposure without acute poisoning and depression was not statistically significant (OR = 1.12; 95% CI, 0.93–1.35, *p* = 0.221), underscoring the need for further longitudinal research to clarify the long-term mental health effects of chronic pesticide exposure. Additional research incorporating biomarkers and socioeconomic factors is essential to better understand the relationship between pesticide exposure and mental health. Organophosphate (OP) insecticides, widely used in forestry and agriculture, remain a significant health concern due to their toxicity.^2^ While high-income countries have tightened pesticide regulations, low- and middle-income countries like Chile continue to use hazardous pesticides,^16^ with residues frequently detected in locally consumed produce, including fruits and vegetables.^17,18^ Moreover, pesticide imports in Chile have increased substantially, doubling between 2005 and 2019.^19^

In recent years, pesticide poisoning cases have risen in Chile. In 2023, the National Epidemiological Surveillance Network for Pesticides (REVEP) reported 724 suspected cases of acute poisoning, 654 of which were confirmed, resulting in an incidence rate of 3.3 per 100,000 inhabitants. This exceeds the 497 cases reported in 2019 and the median of 579 cases expected for the period 2015–2019. Outbreaks were particularly concentrated in the O’Higgins, Maule, and Metropolitan regions.^20^

In the Maule region, organophosphate pesticide metabolites, including methyl parathion, diazinon, and chlorpyrifos, have been detected in the urine of children from rural communities, linked to the proximity of schools to agricultural fields where these pesticides are applied.^21^ A prior study of 190 schoolchildren in the region associated urinary dialkyl phosphate metabolites with fruit and vegetables consumption, proximity to agricultural fields, and pesticide use at home.^22^

Additionally, research in the region has documented symptoms such as peripheral polyneuropathy, anxiety, and reduced neurocognitive performance among farmers exposed to pesticides, although these studies did not incorporate biomarkers to measure exposure.^4^ The Agricultural and Livestock Service^19^ reports that the Maule region ranks second in pesticide sales nationwide (15.7%), following the O’Higgins region (51.2%), both of which are Chile’s primary agricultural areas. Chlorpyrifos and diazinon are the most commonly sold insecticides in the Maule region. Notably, a study in the Coquimbo region found that 60% of agricultural workers possess a genotype that reduces their efficiency in metabolizing chlorpyrifos, heightening their risk of health issues from prolonged exposure.^23^ Although Chile’s Ministry of Agriculture^24^ recently banned chlorpyrifos, the regulation is in transition, allowing its sale and distribution for two years or until existing stocks are depleted.

This study aims to contribute to the literature on pesticide exposure and depressive symptoms by analyzing the association between organophosphate pesticide exposure and depressive symptoms among rural and urban workers in the Maule region, Chile. The study’s relevance lies in enhancing the understanding of mental health risks associated with pesticide exposure across different populations and providing evidence to support the development of more effective policies to protect individuals at potential risk.

## 2. Methods

This longitudinal study aimed to assess the association between organophosphate pesticide exposure and depressive symptoms among agricultural and non-agricultural workers in the Maule region of Chile. The agricultural group, representing the rural population with occupational pesticide exposure, was randomly selected from the database of the Agricultural Development Institute of Chile (INDAP). The non-exposed group, consisting of urban workers without occupational pesticide exposure, was recruited from transportation, sanitation, and construction company databases.

The sample size was calculated based on prior research identifying significant neurophysiological differences in neurobehavioral responses between agricultural workers and a control group.^25^ To account for potential participant attrition, a 20% oversampling was applied, resulting in a final total of 80 participants (n = 40 per group). Participants were matched by age, sex, and educational level to control for confounding variables. During exploratory analysis of DAP metabolite levels, one urban non-occupationally exposed participant exhibited unexpectedly high metabolite concentrations during a period of minimal agricultural activity. Subsequent investigation revealed non-agricultural occupational use of materials and chemicals containing organophosphate-related active compounds, leading to their exclusion from the analysis. Among participants who did not continue in the study during the second year, one rural exposed worker and two urban non-exposed workers withdrew due to health issues.

Additionally, one urban non-exposed worker discontinued participation after experiencing a material loss of their home, and another urban non-exposed worker withdrew voluntarily from the study.

Pesticide exposure was assessed through the average concentrations of six dialkyl phosphate (DAP) metabolites: dimethyl phosphate (DMP), dimethyl thiophosphate (DMTP), dimethyl dithiophosphate (DMDTP), diethyl phosphate (DEP), diethyl thiophosphate (DETP), and diethyl dithiophosphate (DEDTP). To account for the 48-hour metabolism period of pesticides in the human body and enhance measurement accuracy,^21,22^ urine samples were collected twice within the same week (Tuesday and Thursday) during two years (2022 and 2023) (See Figure 1).

**Figure 1.**
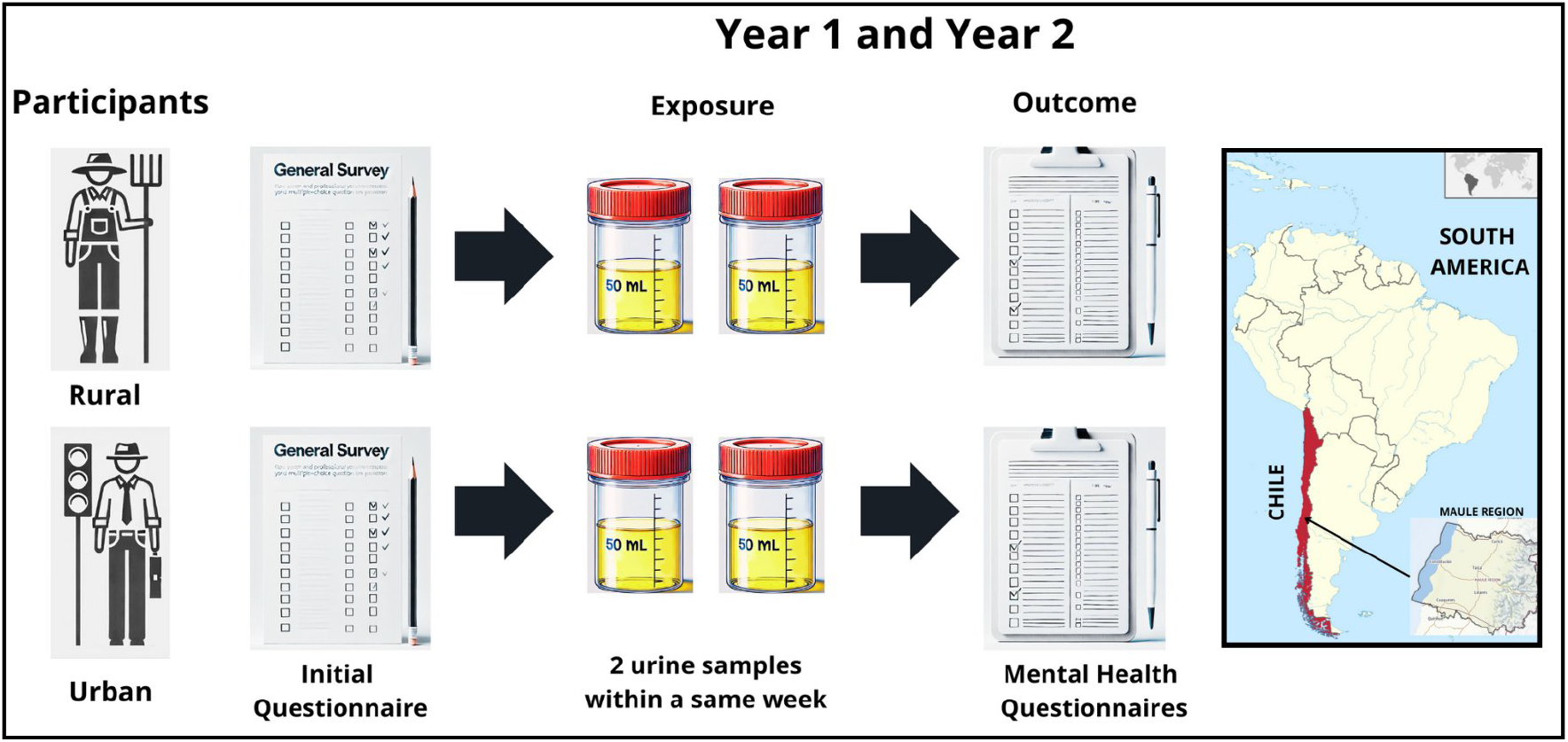
Schematic representation of the basic data collection process and a map showing the location of the Maule Region, where the study was conducted.

Urine samples were collected in 50 mL sterile bottles, frozen at -20°C, and shipped to the Centers for Disease Control and Prevention (CDC) in Atlanta, USA, on dry ice for analysis using tandem mass spectrometry with isotope-dilution quantification. Creatinine levels were measured in each sample using a colorimetric method to adjust metabolite concentrations for each participant. Urine collection and analysis followed established CDC laboratory protocols.^21^ Participants were briefed in advance by the research team and prepared for sample collection at their homes. A summary of the data collection process and the location of the study can be seen in figure 1.

Depressive symptomatology was assessed one to two months following urine sample collection in 2022 and 2023 using the CES-D scale, a validated self-report instrument in Chile ^26^ with high sensitivity (98%) and specificity (79%). The CES-D scale comprises 20 items, generating a total score ranging from 0 to 60, with a cutoff of ≥16 indicating depressive symptoms. Positive and negative affect were evaluated using the PANAS questionnaire, with the Chilean version^27^ demonstrating acceptable reliability (Cronbach’s alpha: 0.529 – 0.768). Low positive affect scores are indicative of depression, whereas high negative affect scores are associated with stress or anxiety (See Figure 1).

At the start of the study, participants completed a comprehensive questionnaire to collect sociodemographic, health, and exposure-related variables.^21^ Data included age, gender, educational level, marital status, monthly household income, family size, physical health history, prior diagnoses of depression and anxiety, proximity of residences (and workplaces for the urban participants) to agricultural fields, duration of current employment, pesticide use at work and/or home, and history of pesticide poisoning.

To evaluate the risk of non-medical substance use, the ASSIST 3.0 questionnaire^28^ was administered. This tool assesses substance use patterns through eight items, including lifetime use, use within the past three months, and associated problems across ten substances, such as tobacco, alcohol, cannabis, and cocaine. Risk levels were categorized as low, moderate, or high, providing a framework for guiding intervention strategies. The administration time ranged from 5 to 15 minutes, depending on the number of substances reported by participants. A summary of the data collection process and the location of the study can be seen in figure 1.

Data analysis began with exploratory analyses, followed by univariate and bivariate statistical tests. Multiple regression analysis was performed to examine the relationship between urinary DAP metabolite concentrations and symptoms of anxiety and depression, controlling for potential confounders such as educational attainment, substance use (drug/alcohol), age, gender, and the number of trials retained in mean waveforms. Data from the two-year study period were combined, and Generalized Estimating Equations (GEE) were used to account for repeated measures across time points.

Urinary DAP concentrations below the limit of detection (LOD) were assigned a value of LOD/√2.^29^ Concentrations were converted to SI units to calculate molar concentrations, and diethyl (DEAP) and dimethyl (DMAP) metabolites were summed to generate aggregated exposure metrics (ΣDEAP and ΣDMAP).^21^ These summed metabolites were treated as continuous variables in the analysis.

Ethical approval for the study was obtained from the Scientific Ethics Committee of Universidad Católica del Maule, Chile (n°16/2022).

## 3. Results

The initial sample in year 1 comprised 78 participants, including 41 occupationally exposed individuals (82 repeated measurements) and 37 non-occupationally exposed individuals (74 repeated measurements).

In year 2, there was a 6% sample loss due to health or employment changes. The final sample consisted of 40 exposed workers (80 repeated measurements) and 33 non-exposed workers (66 repeated measurements). Across the two study years, a total of 306 urine samples were collected and analyzed (Figure 1).

The results summarized in Table 1 compare key characteristics of rural agricultural workers occupationally exposed to pesticides with urban non-agricultural workers not occupationally exposed during year 1 and year 2. The rural and urban workers demonstrated comparable average ages, with a mean of approximately 54–55 years (males were 90% of rural workers and 87% of urban workers). A reliably higher proportion of rural exposed workers had incomplete secondary or primary education compared to urban workers (*p* < 0.05). Furthermore, per capita income was higher among rural workers for both years (*p* < 0.05).

**Table 1.** Demographic and Occupational Characteristics of Rural Agricultural and Urban Non-Agricultural Exposed Workers in year 1 and year 2.

| Variable | Year 1 (2022) |  | Year 2 (2023) |  |
| --- | --- | --- | --- | --- |
|  | Rural<br>n= 41 | Urban<br>n= 37 | Rural<br>n= 40 | Urban<br>n= 33 |
| Age (M, SD) | 54 (10) | 54.6 (8.3) | 55 (9.8) | 55 (8.3) |
| Male (%) | 90% | 87% | 90% | 87% |
| Incomplete secondary education or less (%) | 68% | 9%* | 67% | 16%* |
| Income per capita<sup>a</sup> (M,SD) | \$122,551 (56,649) | \$192,314 (127,059)* | \$132,606 (83,839) | \$208,890 (125,425)* |
| Married or cohabitating (%) | 56% | 68%* | 61% | 71% |
| Alcohol consumption (%) | 56% | 57% | 69% | 81% |
| Tobacco consumption (%) | 10% | 27%* | 18% | 28% |
| Pesticide use at home (%) | 73% | 33%* | 65% | 39%* |
| Chronic disease (self-reported, medical diagnosis) (%) | 66% | 43%* | 72% | 58% |
| Depression (self-reported, medical diagnosis) (%) | 17% | 5%* | 13% | 9% |

\* Mann-Whitney U $p < 0.05$
<sup>a</sup> Values in Chilean pesos. As of the current exchange rate, one US dollar is approximately 980 Chilean pesos.

A higher proportion of urban non-exposed workers were married or living with a partner in year 1 (*p* < 0.05). Alcohol consumption increased in both groups in year 2, with a higher prevalence among urban (81%) compared to rural workers (68%). Tobacco consumption was higher among urban workers in year 1 and 2 (*p* < 0.05). Rural exposed workers reported more frequent pesticide application at home in both years (*p* < 0.05).

Chronic diseases were more prevalent among rural than urban workers in both years, stronger in year 1 (66% vs. 42%). Additionally, the prevalence of self-reported depression diagnoses was significantly higher among rural workers in year 1 compared to urban workers (*p* < 0.05); however, this difference diminished in year 2.

Among agricultural workers, 54% reported applying organophosphate pesticides at home, and 44% indicated infrequent or inconsistent use of personal protective equipment (PPE) during pesticide application. Table 2 provides the distribution of workers applying pesticides by active compound.

**Table 2.** Proportion of Rural Workers Applying Pesticides by Active Compound with high sensitivity (98%) and specificity (79%). The CES-D scale comprises 20 items, generating a total score ranging from 0 to 60, with a cutoff of ≥16 indicating depressive symptoms. Positive and negative affect were evaluated using the PANAS questionnaire, with the Chilean version

| Pesticide | % (n=40) |
| --- | --- |
| Methamidophos | 46% |
| Chlorpyrifos | 40% |
| Methomyl | 19% |
| Lambda Cyhalothrin | 17% |
| Benomyl | 16% |
| Carbofuran | 14% |
| Paraquat | 12% |
| Diazinon | 11% |
| Glyphosate, Mancozeb, Azinphos methyl, Deltamethrin, Oxyfluorfen * | 5% |
| Imidacloprid (Deltamethrin), Pirimicarb, Cadusafos* | 4% |
| Profenophos, Acloniphene * | 3% |
| Fluazifop-p-butyl, Clethodima, Chlorantraniliprole, Imidacloprid, Pendimethalin, Metolachlor, Cypermethrin, Avermectis, Boscalid (Pyraclostrobin), Indoxacarb, Beta Cyfluthrin, Aldicarb* | 1% |

\* The percentages in the right column represent each pesticide listed in the row, grouped for clarity and space efficiency; they are not additive relative to the total % shown.

Table 2 highlights that insecticides are the most commonly used class of pesticides among workers, followed by herbicides and fungicides. Methamidophos, an organophosphate insecticide, was the most frequently reported pesticide, applied by 46% of workers, followed by Chlorpyrifos, another organophosphate insecticide, used by 40% of workers. Additional insecticides included the carbamate Methomyl, reported by 19%, and the pyrethroid Lambda-Cyhalothrin, reported by 17% of workers. Among herbicides, Glyphosate and Paraquat were the most commonly used, reported by 5% and 12% of workers, respectively. For fungicides, Benomyl was the most frequently applied, used by 16% of workers.

### 3.1. Differential metabolic processing of pesticides between rural and urban workers as evidenced by diethyl and dimethyl alkylphosphates concentrations in urine

The analysis of dialkyl phosphate (DAP) metabolites showed that median concentrations of diethyl alkylphosphates (ΣDEAP) were consistent across the two years (19.90 [11.13–35.91] in 2022 vs. 18.81 [10.54–36.79] in 2023), with no reliable differences (*p* = 0.9867), indicating stability in this metabolite over time. In contrast, dimethyl alkylphosphates (ΣDMAP) showed a clear reduction in the median concentration from 31.46 [19.67–61.67] in year 1 to 19.97 [10.09–34.42] in year 2 (*p* < 0.0001), with increased variability and higher maximum values observed in year 2.

The total DAP concentrations (ΣDAP) also reliably decreased, with a median of 45.42 [29.12–90.33] in year 2 compared to 56.31 [36.68–96.90] in year 1 (*p* = 0.0492). These findings suggest changes in exposure sources, pesticide application practices, or metabolic factors influencing DAP levels over time. The increased variability and extreme values in year 2 highlight the need for further investigation into potential temporal trends and their implications for health outcomes.

Table 3 summarizes the median concentrations and interquartile ranges (IQR) of dialkyl phosphate (DAP) metabolites among rural agricultural workers and urban non-agricultural workers for year 1 and year 2. Rural agricultural workers exhibited higher concentrations of diethylthiophosphate (DETP) compared to urban workers, stronger for year 2. This increase may reflect heightened occupational exposure associated with specific agricultural practices or conditions; these differences, however, were not consistent across all metabolites or study years, underscoring significant individual variability within groups, as indicated by the wide IQRs.

**Table 3.** Concentrations of dialkyl phosphates (DAPs) among rural agricultural workers (occupationally exposed) and urban non-agricultural workers (non-occupationally exposed) in Year 1 and Year 2.

| Metabolites | Year 1(2022) |  | Year 2(2023) |  |
| --- | --- | --- | --- | --- |
|  | Rural<br>Median (IR) | Urban<br>Median (IR) | Rural<br>Median (IR) | Urban<br>Median (IR) |
| DEP $\mu\text{g/L}$ | 2.00 (0.98-5.12) | 2.18 (1.06-4.08) | 2.22 (1.18-6.58) | 1.80 (0.98-3.75) |
| DEP $\mu\text{g/g cr}^a$ | 2.22 (1.15-3.88) | 2.84 (1.77-6.16) | 2.33 (1.38-6.46) | 2.55 (1.45-3.98) |
| DEP nmol/L<sup>a</sup> | 14.47 (7.45-25.16) | 18.48 (11.53-40.06) | 15.11 (8.9-41.93) | 16.62 (9.46-25.86) |
| DETP $\mu\text{g/L}^b$ | 0.42 (0.21-0.75) | 0.27 (0.12-0.64) | 0.39 (0.21-1.44) | 0.27 (0.13-0.53) |
| DETP $\mu\text{g/g cr}$ | 0.42 (0.26-0.84) | 0.39 (0.20-0.79) | 0.35 (0.21-1.30) | 0.35 (0.21-0.57) |
| DETP nmol/L | 2.45 (1.53-4.94) | 2.33 (1.18-4.68) | 2.09 (1.27-7.69) | 2.11 (1.27-3.39) |
| ΣDAPs nmol/L<sup>a</sup> | 17.40 (9.10-32.15) | 23.50 (13.06-43-34) | 15.11 (8.91-41.92) | 16.62 (9.45-25.86) |
| DMP $\mu\text{g/L}$ | 3.71 (1.63-7.92) | 3.12 (1.10-5.39) | 2.29 (0.82-6.00) | 1.54 (0.90-3.6) |
| DMP $\mu\text{g/g cr}^a$ | 3.50 (2.07-5.76) | 3.75 (2.28-6.99) | 1.81 (0.78-4.81) | 2.44 (1.12-3.33) |
| DMP nmol/L | 27.80 (16.45-45.77) | 29.83 (18.11-55.49) | 1.81 (1.11-4.81) | 2.44 (1.12-3.33) |
| DMTP $\mu\text{g/L}$ | 2.08 (1.2-4.57) | 2.13 (0.87-5.27) | 1.52 (0.57-3.44) | 1.16 (0.51-2.5) |
| DMTP $\mu\text{g/g cr}^a$ | 0.24 (0.15-0.33) | 0.32 (0.14-0.69) | 14.39 (8.78-38.16) | 19.38 (8.94-26.46) |
| DMTP nmol/L<sup>a</sup> | 1.70 (1.02-2.31) | 2.28 (1.04-4.89) | 1.02 (0.59-1.67) | 1.29 (0.66-1.97) |
| DMDTP $\mu\text{g/L}$ | 0.19 (0.11-0.36) | 0.26 (0.07-0.53) | 0.13 (0.07-0.24) | 0.12 (0.07-0.21) |
| DMDTP $\mu\text{g/g cr}^a$ | 0.24 (0.15-0.33) | 0.32 (0.14-0.69) | 0.15 (0.08-0.24) | 0.18 (0.09-0.28) |
| DMDTP nmol/L<sup>a</sup> | 1.53 (0.92-2.10) | 2.04 (0.94-4.40) | 0.92 (0.53-1.51) | 1.16 (0.59-1.77) |
| ΣDMAPs nmol/L | 30.92 (18.70-50.00) | 34.52 (21.65-63.04) | 17.30 (10.01-45.36) | 21.92 (11.00-31.37) |
| ΣDAPs nmol/L | 52.52 (35.59-78.91) | 67.35 (42.08-102.43) | 55.10 (26.32-125.72) | 42.52 (30.79-63.68) |
| Creatinine g/L<sup>a</sup> | 1.11 (0.68-1.41) | 0.76 (0.51-1.18) | 0.99 (0.67-1.38) | 0.85 (0.50-1.39) |

DEP: Diethylphosphate; DETP: Diethylthiophosphate; DMP: Dimethylphosphate; DMTP: Dimethylthiophosphate; DMDTP: Dimethyldithiophosphate; ΣDEAPs: Sum of diethyl alkylphosphates; ΣDMAPs: Sum of dimethyl alkylphosphates; $\Sigma$ DAPs: Total sum of dialkyl phosphates. The DAP concentrations were converted to their molar equivalents and then added up to determine the composite concentrations.
<sup>a</sup> $p < 0.05$ U Mann Whitney year 1; <sup>b</sup> $p < 0.05$ U Mann Whitney year 2

In contrast, urban non-agricultural workers exhibited higher concentrations of certain metabolites in year 1, including the sum of diethylalkylphosphates (ΣDEAPs) and dimethylalkylphosphates (ΣDMAPs). This finding may reflect potential environmental exposure in non-occupational settings or the ingestion of pesticide residues through food. Creatinine-normalized concentrations further indicated that urban workers had comparatively higher values in specific instances, suggesting physiological or environmental factors that could influence the excretion patterns of these compounds.

### 3.2. Increased depressive scores for rural workers

Rural agricultural workers exhibited slightly higher median CES-D scores for depressive symptomatology in both Year 1 (21.5 (13-30) vs. 19.0 (13-24)) and Year 2 (23.1 (17-30.5) vs. 21 (15-26)) than their urban counterparts; however, these differences were not reliably significant. In Year 2 (measures 3 and 4) , both rural and urban groups reported higher depressive scores compared to Year 1, potentially indicating a link to increased environmental exposure (Figure 2).

**Figure 2.**
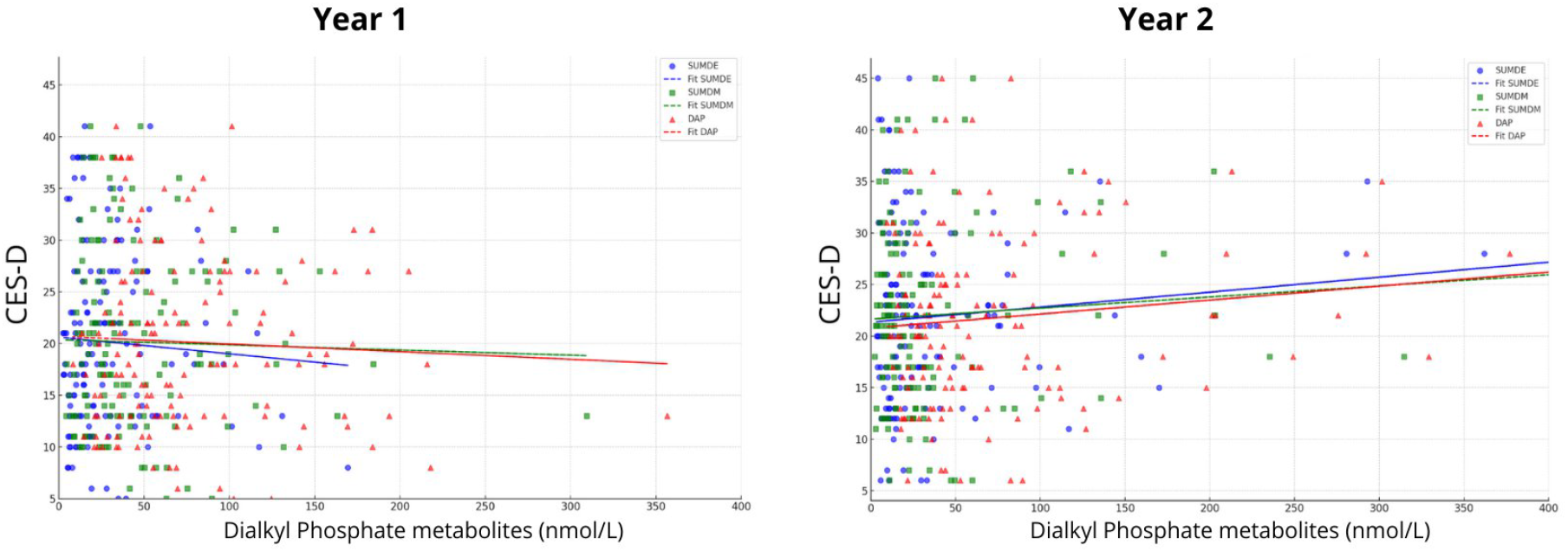
Scatterplots illustrating the associations between the concentration levels (nmol/L) of each sum of Dialkyl Phosphate metabolites (DEAPs, DMAPs, and DAPs) and CES-D scores (a measure of depressive symptoms). Data from both rural and urban participants are included across all measurement time points (Year 1: Timepoints 1 and 2; Year 2: Timepoints 3 and 4). To enhance visualization, the x-axis is restricted to a range of 0–400 nmol/L; however, all data points, including outliers, are included in the estimation of the linear regression lines. The graphs indicate stronger associations with CES-D scores during the second year, particularly for DEAPs.

Regarding positive affect, as measured by the PANAS scale, urban workers exhibited significantly higher median scores compared to rural workers in Year 1 (34.0 (31-37.5) vs. 38.0 (35-41) (p < 0.05). However, this trend reversed in Year 2, (39.3 (35-43.5) vs. 34.8 (32-39)). This shift may reflect the temporal effects of exposure on positive emotional states. For negative affect, rural workers consistently exhibited higher median scores in both years. In Year 2, urban workers showed a significant reduction in median negative affect compared to rural workers (21.3 (16.5-27) vs. 16.8 (10-20) (p < 0.05). These findings suggest a potential association between occupational pesticide exposure and elevated negative affect, reflecting heightened anxiety among exposed workers. The concurrent presence of high negative affect and elevated positive affect in exposed workers may reflect an ambivalent emotional response, wherein the stress of prolonged exposure coexists with efforts to maintain emotional well-being through coping mechanisms.^30^

### 3.3. Cumulative exposure to diethyl alkyl phosphates (DEAPs) predicts depressive symptoms

A dose-response analysis was performed using generalized estimating equation (GEE) models to evaluate the cumulative effect of pesticide exposure over the two-year period. In developing the final model, sociodemographic variables that demonstrated significant differences between rural and urban groups in bivariate analyses were initially included. Subsequently, variables identified in the literature as being associated with depressive symptomatology—such as alcohol consumption, previous depression diagnosis, and sex—were incorporated. To enhance interpretability, the model was further adjusted for creatinine levels to account for variability in urinary excretion, given that pesticide exposure was measured through dialkyl phosphate metabolites. Sociodemographic and health covariates with a p-value < 0.1 were retained in the final model.

In the GEE analysis (Table 5), only the sum of DEAP metabolites in urine and CES-D depressive symptomatology showed a significant association (Wald χ^2^ = 30.41; p = 0.0001).

**Table 5.** Results of the Generalized Estimating Equations (GEE) model assessing the association between exposure <u>to diethylphosphate pesticides(DEAPs) and depressive symptomatology, adjusted for covariates (2022–2023).</u>.

| Variable | Coefficient | SE | z | p-value | 95% CI |  |
| --- | --- | --- | --- | --- | --- | --- |
| Sum of DEAPs (nmol/L)* | 0.0065 | 0.0031 | 2.06 | 0.040 | 0.0003 | 0.0127 |
| Gender | -5.8229 | 2.5920 | -2.25 | 0.025 | -10.90 | -0.742 |
| Chronic disease | 2.0004 | 1.1307 | -1.77 | 0.077 | -0.215 | 4.216 |
| Depression (diagnosis) | 5.2452 | 1.6558 | 3.17 | 0.002 | 1.999 | 8.490 |
| Problematic alcohol consumption | 1.8630 | 0.9409 | 1.98 | 0.048 | 0.018 | 3.707 |

\*Creatinine adjusted

The coefficient for cumulative pesticide exposure (β = 0.0065, p = 0.040) indicates a dose-response relationship, with higher exposure levels associated with increased severity of depressive symptoms. Additionally, and unsurprisingly, individuals with a history of depression exhibited significantly higher CES-D scores (β = 5.2452, p = 0.002), aligning with evidence supporting the recurrent nature of depressive disorders.^31^ Problematic alcohol consumption was also positively associated with depressive symptoms (β = 1.8630, p = 0.048), corroborating previous research highlighting the strong connection between substance use and mental health disorders.^32^

Sex emerged as a significant predictor (β = -5.8229, p = 0.025), indicating that men reported fewer depressive symptoms compared to women, consistent with established literature on gender differences in mental health outcomes although the interpretability of this is low as the gender proportion was ∼90/10. Although chronic disease (p = 0.077) and per capita income were not statistically significant in this model, they remain important factors for understanding variability in depressive symptoms. These findings underscore the need to address cumulative pesticide exposure and behavioral factors, such as alcohol use, in efforts to reduce the burden of depressive symptoms in affected populations.

## 4. Discussion

This longitudinal study examined the relationship between organophosphate pesticide exposure and depressive symptoms among rural and urban workers in the Maule region of Chile. The findings indicated that increased pesticide exposure, particularly as measured by the sum of diethyl alkyl phosphate (DEAP) metabolites, was reliably associated with a rise in depressive symptoms. These results are consistent with previous studies that have identified a link between pesticide exposure and the development of mental health disorders, including depression.^3,8,9,33^ A possible explanation for why more associations were observed with diethyl (DEAP) metabolites than dimethyl (DMAP) metabolites may lie in the chemical and metabolic properties of these compounds and their differing biological processes in the body. Diethyl organophosphates, which include fat-soluble compounds, are metabolized into molecules that may persist longer in the body. Their fat solubility allows them to remain in circulation at low levels over extended periods, potentially leading to more prolonged or cumulative neurotoxic effects compared to dimethyl organophosphates.^34,35^ These findings are consistent with prior research that highlights the varying neurotoxic potentials of different organophosphate subgroups that produce differential effects.^36,37^ Future studies should aim to elucidate the specific biochemical mechanisms through which diethyl organophosphates may contribute to depressive symptoms, including their interactions with neurotransmitter systems and inflammatory pathways. Investigating genetic and epigenetic factors that modulate individual susceptibility to these compounds may also provide valuable insights.^23^

The dose-response relationship observed in this study reinforces existing evidence on the mental health risks associated with pesticide exposure. It should be noted that the effects found in a relatively small sample highlight the strength and variability of the exposure and are a cautionary tale for the responsible use and regulation of these pesticides, at the industry, government and societal level. These results align with the meta-analysis by Frengidou et al.,^3^ which reported that acute pesticide poisoning nearly triples the risk of depression compared to non-poisoned individuals. However, this study offers a novel contribution by employing specific biomarkers, such as DAPs metabolites, to evaluate the relationship between cumulative pesticide exposure and depressive symptoms—an approach that has been underexplored in Latin American research.

Unlike previous studies that primarily focused on acute pesticide exposure,^13,14^ this study emphasizes chronic exposure and its association with depressive symptoms, highlighting variability and cementing the cumulative effects. Although research such as Cancino et al.^8^ suggests that chronic pesticide exposure may elevate the risk of depression, the evidence remains inconclusive. This study contributes to the ongoing discussion by demonstrating that chronic and cumulative pesticide exposure, as assessed through biomarkers, may be associated with increased depressive symptoms, even in the absence of acute poisoning.

Regarding problematic alcohol consumption, individuals with problematic alcohol use exhibited a higher risk of depressive symptoms, consistent with studies such as Castillo-Carniglia et al.,^32^ which identified a bidirectional relationship between substance use and depression. These results highlight the importance of considering alcohol consumption habits when evaluating mental health in populations exposed to pesticides, as alcohol use may exacerbate depressive symptoms. Conversely, chronic use of alcohol may interfere with the efficiency of the metabolic pathways processing the organophosphates.

Further societal parameters, such as gender, per capita income, and creatinine levels did not show significant associations with depressive symptoms in our model. While existing literature suggests that gender may influence the manifestation of depressive symptoms,^31^ no significant differences were observed in our study population, with the caveat of a highly biased sample. Regarding per capita income, although previous research has indicated that socioeconomic conditions can affect mental health,^8^ no clear association was identified in this study. This may be attributed to the relative socioeconomic homogeneity of our sample, which predominantly comprised low-income workers. Adjusting for creatinine levels was a critical aspect of this study, as it controlled for variability in urinary excretion, ensuring that differences in pesticide metabolite levels were attributable to exposure rather than physiological variations in substance elimination. This adjustment enhances the accuracy of pesticide exposure interpretation, aligning with recommendations from previous studies utilizing exposure biomarkers.^21^

One of the key strengths of this study is its longitudinal design and the use of specific biomarkers to measure pesticide exposure, enabling a more precise assessment of the relationship between cumulative exposure and depressive symptoms. Furthermore, the collection of urine samples at two time points during the peak pesticide application period enhances the validity of our findings by effectively capturing exposure during critical periods. However, the study also has certain limitations. The relatively small sample size may have constrained the analysis’ ability to detect more subtle associations, particularly for variables such as income or gender. Additionally, the reliance on self-reported data for variables like depression diagnosis and alcohol consumption could have introduced recall bias or underreporting. A more data-driven analysis might reveal hidden or latent variables to complement the direct approach taken in this study.^38^

A novel aspect of this study is the inclusion of an urban non-occupationally exposed sample of the population, enabling a comparison of the effects of indirect pesticide exposure among non-agricultural workers. While these individuals do not directly handle or apply pesticides, they experience environmental exposure due to living in a region with agricultural fields where pesticides are used and high agricultural activity.^39^ This underscores the importance of addressing both occupational and environmental exposure, particularly during peak pesticide application periods, as observed in year 2(2023). Despite being indirectly exposed, non-occupational populations may still face a risk of adverse health effects, highlighting the need for further investigation in future studies.

A novel aspect of this study is the inclusion of an urban, non-occupationally exposed sample of the population, enabling a comparison of the effects of indirect pesticide exposure among non-agricultural workers. These individuals, although not directly handling or applying pesticides, live in the Maule Region of Chile, a location distinguished by its extensive agricultural fields and intense agricultural activity.^39^ This highlights the critical importance of addressing both occupational and environmental pesticide exposures, especially during peak application periods, as observed in year 2 (2023). Even individuals with indirect exposure may face significant health risks, demonstrating that pesticide exposure is not solely an occupational issue but a broader public health concern in regions with high agricultural activity.

Based on our findings, we recommend that future research include larger sample sizes and longer follow-up periods to better evaluate the long-term effects of pesticide exposure on mental health. Additionally, as suggested by studies conducted in other regions,^23^ investigating genetic factors that may influence susceptibility to depressive symptoms following pesticide exposure, could provide valuable insights. We also urge decision-makers in similar occupational contexts to implement stricter policies aimed at reducing both occupational and environmental pesticide exposure and to promote the consistent use of personal protective equipment among agricultural workers. ^4,40,41^ Furthermore, it is essential to address environmental exposure in communities located in regions with extensive agricultural fields and to develop targeted strategies to mitigate the associated mental health risks in these populations.^42^ Thus, these findings provide valuable input for policymakers to establish surveillance protocols that encompass not only farmworkers but entire communities, ensuring comprehensive monitoring and effective mitigation of agrochemical exposure risks.

Our findings also suggest several important recommendations for clinical and research practices. In clinical settings, it would be valuable to include a question about pesticide use or exposure during the anamnesis of patients presenting with depression. This is particularly important for individuals who may be chronically exposed to pesticides, such as those with occupational exposure. Identifying such exposure could inform a more comprehensive treatment strategy, as continued exposure to pesticides might exacerbate depressive symptoms and undermine the efficacy of therapeutic interventions. Addressing the source of exposure alongside treatment could prevent a cycle of ineffective care where patients return to conditions that perpetuate their risk of depression.

For future research, exploring the interaction between pharmacological treatments for depression and pesticide exposure is a critical avenue. Understanding how antidepressant drugs interact with pesticides at a biological level could help mitigate iatrogenic risks, such as adverse reactions stemming from the combined effects of these chemicals. Such studies would enhance our ability to design safer and more effective therapeutic protocols for individuals exposed to pesticides, ultimately improving both mental health outcomes and occupational safety standards. These recommendations highlight the need for a multidisciplinary approach that integrates environmental, occupational, and mental health considerations in both clinical practice and research.

In conclusion, this study presents compelling evidence of an association between pesticide exposure and depressive symptoms in agricultural workers, underscoring the importance of addressing both occupational and environmental exposures. The findings highlight the necessity of developing more effective public health policies to safeguard vulnerable populations and implementing comprehensive surveillance protocols that extend to entire communities. Future research is suggested to investigate the mechanisms underlying these effects and to examine the interaction between pesticide exposure and depression treatments, with the aim of informing safer therapeutic strategies and advancing the understanding of the long-term impacts of pesticide exposure on mental health.

## Data Availability

All data produced in the present study are available upon reasonable request to the author

